# Molecular diagnosis of non-syndromic hearing loss patients using a stepwise approach

**DOI:** 10.1101/2020.09.20.20197145

**Authors:** Jing Wang, Jiale Xiang, Lisha Chen, Hongyu Luo, Xiuhua Xu, Nan Li, Chunming Cui, Jingjing Xu, Nana Song, Jiguang Peng, Zhiyu Peng

## Abstract

**Background:** Hearing loss is one of the most common birth disorders in humans with an estimated prevalence of 1-3 in every 1000 newborns. This study has investigated the molecular etiology of a deaf cohort using a stepwise strategy to effectively diagnose patients and the challenges faced to verify genetic heterogenicity and the variable mutation spectrums of hearing loss.

**Methods:** In order to target known pathogenic variants, multiplex PCR plus next-generation sequencing was applied in the first tier, while undiagnosed cases were further referred to exome sequencing. A total of 92 unrelated patients with nonsyndromic hearing loss were enrolled.

**Results:** In total, 64% (59/92) of patients were molecularly diagnosed, 44 of which were identified in the first tier by multiplex PCR plus sequencing. Of 48 undiagnosed patients from the first tier, exome sequencing identified eleven diagnoses (23%, 11/48) and four probably diagnoses (8%, 4/48). The rate of secondary findings of exome sequencing in our cohort is 3.4%.

**Conclusion:** The research presented a molecular diagnosis spectrum of 92 non-syndromic hearing loss patients and demonstrated the benefits of using the stepwise diagnostic approach in the genetic test of the non-syndromic hearing loss patient cohort.

## INTRODUCTION

Hearing loss is one of the most common birth disorders in humans with an estimated prevalence of 1-3 in every 1000 newborns.^1^ Seventy percent of hearing loss cases are nonsyndromic, and one of the primary etiologies is attributable to genetic predisposition.^1 2^ As on date, over 100 genes have been associated with nonsyndromic hearing loss (NSHL),^3 4^ and new genes are being discovered with time.^5^ The timely and effective diagnosis of affected individuals is challenged by the genetic heterogenicity.

Despite the high heterogenicity, the most common mutations found in distinctive populations are from the *GJB2* gene, which encodes the connexin 26 protein and causes severe-to-profound autosomal recessive nonsyndromic hearing loss.^2 6^ Therefore, *GJB2* Sanger sequencing was first performed in previous studies.^7 8^ Interestingly, the frequency of causative genes shows a variation across different populations and ethnicities. The OTOF was revealed to be a major and potential contributor to hearing loss, instead of *GJB2*, in the Saudi population.^8^ The *SLC26A4* is another preliminary gene causing nonsyndromic hearing impairment with enlarged vestibular aqueduct in Asian, Middle Eastern and Ashkenazi Jews,^9^ while mutations in *GJB2* accounts for 21% of congenital hearing loss.^10^ A single Sanger sequencing was capable of covering the whole region at an affordable cost as the *GJB2* coding region has only 226 amino acids. The *SLC26A4* has 21 exons and the coding sequencing spanned 2343bp from exon 2 to exon 21, which requires multiple Sanger tests.

Although exome sequencing has been proposed and used in hearing loss patients as first tier, ^11 12^ interpreting WES data is usually laborious and time-consuming. Tiered or stepwise diagnose approaches for these patients have been propounded in literature multiple times.^5 7 13 14^ Guan et al. provided a two-tier strategy including combined Sanger and targeted deletion analyses of *GJB2* and *STRC* and two mitochondrial genes, followed by exome sequencing and target analysis of deafness-related genes.^7^ Li et al. performed hotspot variant screening and exome sequencing subsequently in one family with deafness.^5^

This study proposed a hierarchical approach that firstly targets known pathogenic variants by multiplex PCR, followed by exome sequencing and comprehensive analysis of deafness genes in a cohort of 92 non-syndromic hearing loss patients. Exome sequencing was referred to individuals with inconclusive or negative results in the first tier. An analysis of the diagnostic rate in different tiers and the contribution of different genetic factors resulted in a cost-effective diagnostic paradigm, providing an example to other populations.

## MATERIALS AND METHODS

### Participants

A total of 92 patients with nonsyndromic hearing loss were recruited. We had obtained Signed consent from patients. This study was approved by the Institutional Review Board of BGI.

### Multiplex PCR

Genetic mutation detection in all patients was carried out by applying Multiplex PCR combined with next-generation sequencing. The commercial multiplex PCR kit (BGI Biotech, Wuhan, China) was designed to cover certain pathogenic variants of 22 genes, including complete coding region of *GJB2* and most coding regions of *SLC26A4*. Genomic DNA was extracted from 2 ml of peripheral blood by using DNA Extraction Kit (BGI Biotech, Wuhan, China). Targeted variants were amplified by multiplex PCR using 2×KAPA 2G Fast Multiple PCR Mix (KAPA BIOSYSTEMS, Wilmington, MA, USA). The PCR program consists of one round of 95□ for 3min, then 30 cycles including 95□ for 15s, 62□ for 30 seconds, and 72□ for 90s.

### Library preparation, sequencing and bioinformatics

PCR products were pooled to prepare for library. Briefly, ∼3.5ug purified products were sheared by ultrasonoscope and quality-controlled by Agilent Bioanalyzer DNA 2100 kit (Agilent, Santa Clara, CA, USA). Subsequently, the end-repair and A-tailing were performed before adapters were ligated to both ends of the fragments. Finally, the adapter-ligated products were amplified by 8-cycle PCR and purified using Agencourt AMPure XP beads (Beckman Coulter, Fullerton, CA, USA). The prepared libraries were subjected to single-strand circularized DNA and DNA nanoballs before being sequenced on the BGISEQ-500 sequencer (BGI, Shenzhen, China) with PE50.^15^ Raw sequence reads were mapped to the human reference genome (hg19) using the Bowtie 2.3.3 and using SAMtools 1.6 to create BAM and index files. For variant calling, Genome Analysis Tool Kit (GATK 3.7)^16^ was employed to the alignment data and later subjected to a strategic procedure.

### Exome sequencing and data analysis

Genetically undiagnosed patients by multiplex PCR assay were referred to exome sequencing which was performed following standard manufacturer protocols of BGISEQ-500 platform. An in-house bioinformatics pipeline was employed to process variant call format (VCF) files. To maintain variants of potential clinical usefulness, including (i) variants with minor allele frequency (MAF) <1%, (ii) variants in genes with an OMIM disease entry. Consequently, we interpreted variants in 130 genes curated by ClinGen Expert with a limited-to-definitive relationship to hearing loss^17^ were interpreted. This interpretation was based on ClinGen Expert Specification of the ACMG/AMP Variant Interpretation Guidelines for Genetic Hearing Loss.^18^

### Definition of molecular diagnosis

Patients were then categorized as “positive” or “diagnosed” with homozygous or double heterozygous of pathogenic/likely pathogenic variant(s) in a recessive inherited gene, or heterozygous of pathogenic/likely pathogenic variant in a dominant inherited gene. Also, patients with a pathogenic/likely pathogenic variant plus a rare VUS in a recessive inherited gene were titled “probably diagnosed.” Patients with a pathogenic/likely pathogenic variant in a recessive inherited gene were defined as “inconclusive”.

### Sanger validation and qPCR

Sanger sequencing was initiated to validate SNPs/Indels detected by either the multiplex PCR or exome sequencing. All PCR products were sequenced on ABI 3730XL DNA Analyzer. Confirmed mutations were comparing sequencing data to UCSC human reference sequences.^19^ Verification of Exon level deletions or duplications called by exome sequencing was carried out by qPCR.

## Results

Of the 92 non-syndromic hearing loss patients, most (82%; 75/92) were not reported with hearing loss family history. The degree of hearing loss varied; severe-to-profound hearing loss was observed in the majority (84%, 77/92); prelingual hearing loss was detected in 87% (80/92) patients. Of note, 17% (16/92) of patients passed the newborn hearing screens at birth but developed hearing loss in the later stage (Table 1).

**Table 1.** Characteristics of the study cohort ^a^.

| Characteristic | No. (%) |
| --- | --- |
| All | 92 (100) |
| Sex |  |
| Male | 54 (59) |
| Female | 38 (41) |
| Family history |  |
| Yes | 17 (18) |
| No | 75 (82) |
| Onset |  |
| Prelingual ($\leq 3$ y) | 80 (87) |
| Post-lingual ($> 3$ y) | 12 (13) |
| Laterality |  |
| Bilateral symmetric | 69 (75) |
| Bilateral asymmetric | 17 (18) |
| No record | 6 (7) |
| Stability |  |
| Stable | 58 (63) |
| Fluctuating | 24 (26) |
| No record | 10 (11) |
| Aminoglycoside Exposure |  |
| Yes | 4 (4) |
| No | 63 (68) |
| Uncertain | 25 (27) |
| Severity <sup>b</sup> |  |
| Mild | 1 (1) |
| Moderate | 9 (10) |
| Severe | 20 (22) |
| Profound | 57 (62) |
| No record | 5 (5) |
| Rehabilitation |  |
| Hearing aid | 41 (45) |
| Cochlear Implantation | 17 (18) |
| Both | 22 (24) |
| Not applied | 12 (13) |
| Newborn hearing screening result |  |
| Pass | 16 (17) |
| Referral | 36 (39) |
| Not applied / No record | 40 (43) |

a, All members in this cohort have bilateral hearing loss; the precise type of hearing loss are not always recorded, most recorded cases are sensorineural; b, Severity is determined by the hearing level of the better ear; WHO grading rule is adopted (Mild: 26-40dB; Moderate: 41-60dB; Severe: 61-80dB; Profound: $> 80$ dB);

### Forty-four diagnoses by multiplex PCR

In the first step, 44 patients were genetically positive when tested using multiplex PCR, eight inconclusive and 40 negative (Figure 1). The genotypes of 44 patients were listed in table 2, including 27 in *GJB2*, 16 in *SLC26A4*, and 1 in *MT-RNR1*. The index patient who was positive with m.1555A>G in the *MT-RNR1* gene had aminoglycoside exposure history. The homozygote of NM_004004.6:c.235delC in the *GJB2* gene was the most prevalent genotype, accounting 11% (10/92) of the study cohort. Of the 44 patients with positive genotypes, NM_004004.6:c.109G>A in the *GJB2* gene was identified in 10 patients, including three patients with homozygous state and seven patients with compound heterozygous state, respectively. One patient was homozygous for both NM_000441.2:c.919-2A>G in the *SLC26A4* gene and NM_004004.6:c.109G>A in the *GJB2* gene. Considering clinical diagnosis with enlarged vestibular aqueduct, a phenotype which is highly specific for *SLC26A4*,^18^ we thus conclude that NM_000441.2:c.919-2A>G is the causing mutation for this patient.

**Table 2.** Genotype of non-syndromic hearing loss patients detected by Multiplex PCR sequencing.

| No. | Gene | Transcript | Genotype | Inheritance | Number of patients |
| --- | --- | --- | --- | --- | --- |
| 1 | *GJB2* | NM_004004.5 | c.235delC/c.235delC | AR | 10 |
| 2 | *GJB2* | NM_004004.5 | c.235delC/c.299_300delAT | AR | 3 |
| 3 | *GJB2* | NM_004004.5 | c.235delC/c.176_191del | AR | 3 |
| 4 | *GJB2* | NM_004004.5 | c.235delC/c.109G>A | AR | 3 |
| 5 | *GJB2* | NM_004004.5 | c.109G>A /c.109G>A | AR | 3 |
| 6 | *SLC26A4* | NM_000441.1 | c.919-2A>G/ c.919-2A>G | AR | 3* |
| 7 | *GJB2* | NM_004004.5 | c.176_191del/c. 299_300delAT | AR | 1 |
| 8 | *GJB2* | NM_004004.5 | c.299_300delAT/c.109G>A | AR | 1 |
| 9 | *GJB2* | NM_004004.5 | c.109G>A/c.427C>T | AR | 1 |
| 10 | *GJB2* | NM_004004.5 | c.109G>A/c.428G>A | AR | 1 |
| 11 | *GJB2* | NM_004004.5 | c.109G>A/c.583A>G | AR | 1 |
| 12 | *SLC26A4* | NM_000441.1 | c.919-2A>G/c.1174A>T | AR | 1 |
| 13 | *SLC26A4* | NM_000441.1 | c.1226G>A/c.2000T>C | AR | 1 |
| 14 | *SLC26A4* | NM_000441.1 | c.1229C>T/c.1343C>A | AR | 1 |
| 15 | *SLC26A4* | NM_000441.1 | c.1229C>T/c.1692dupA | AR | 1 |
| 16 | *SLC26A4* | NM_000441.1 | c.1229C>T/c.1707+5G>A | AR | 1 |
| 17 | *SLC26A4* | NM_000441.1 | c.1229C>T/c.919-2A>G | AR | 1 |
| 18 | *SLC26A4* | NM_000441.1 | c.1336C>T/c.919-2A>G | AR | 1 |
| 19 | *SLC26A4* | NM_000441.1 | c.1343C>T/c.2168A>G | AR | 1 |
| 20 | *SLC26A4* | NM_000441.1 | c.2000T>C/c.2168A>G | AR | 1 |
| 21 | *SLC26A4* | NM_000441.1 | c.589G>A/c.2168A>G | AR | 1 |
| 22 | *SLC26A4* | NM_000441.1 | c.919-2A>G/c.1991C>T | AR | 1 |
| 23 | *SLC26A4* | NM_000441.1 | c.919-2A>G/c.1614+1G>A | AR | 1 |
| 24 | *SLC26A4* | NM_000441.1 | c.919-2A>G/c.668T>C | AR | 1 |
| 25 | *MT-RNR1* | - | m.1555A>G | Mi | 1 |

\*One patient is homozygous for both *SLC26A4* c.919-2A>G and *GJB2* c.109G>A; AR, autosomal recessive; AD, autosomal dominant; Mi, Mitochondrial

**Figure 1.**
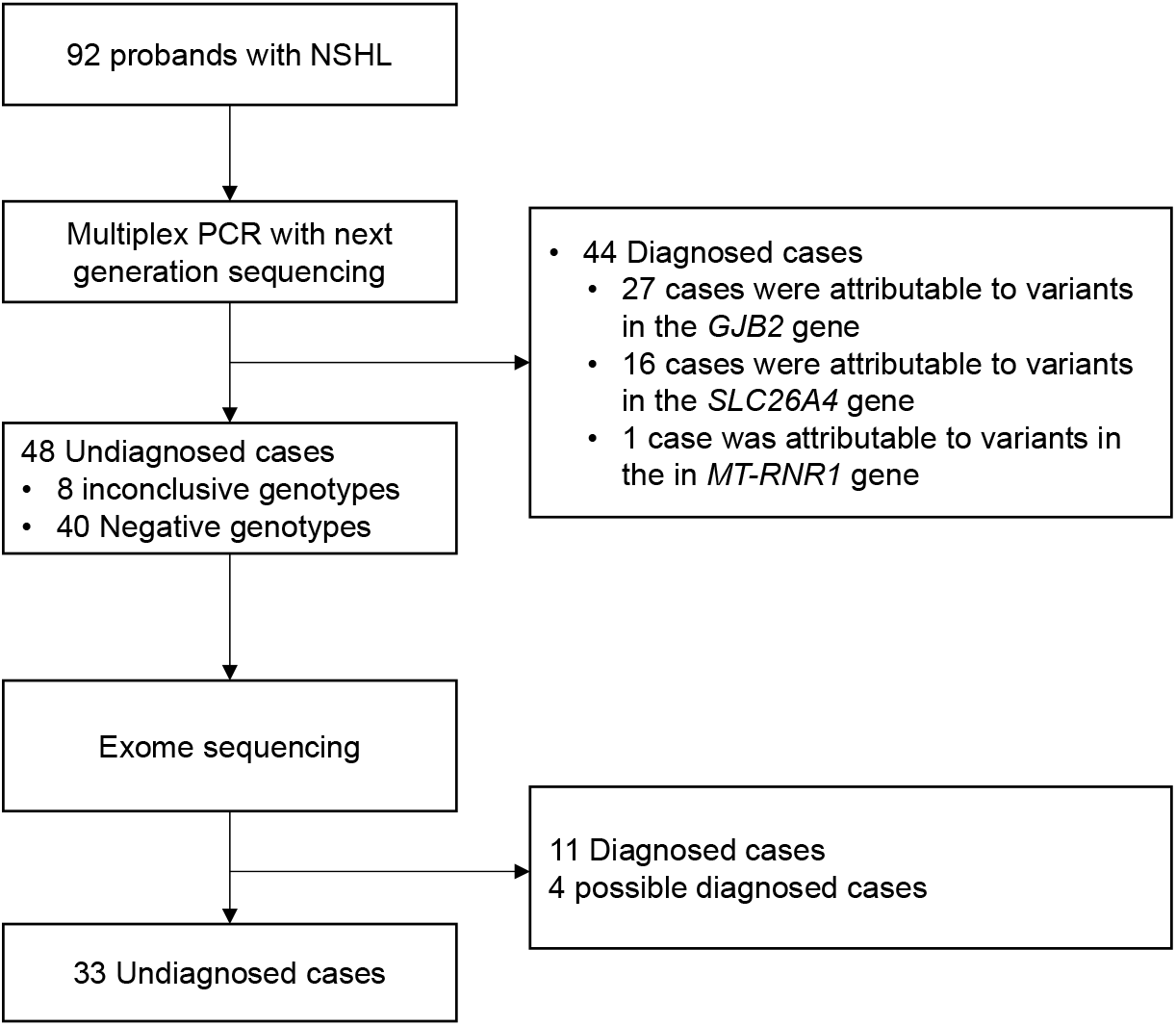
Outline of the study design. Patients suffered from nonsyndromic hearing loss were enrolled. A total of 92 patients were recruited in this study cohort. After performing of Multiplex PCR plus next generation sequencing on all the patients, the 48 undiagnosed and 10 patients diagnosed by *GJB2* c.109G>A were referred to exome sequencing.

### Fifteen diagnoses/probably diagnoses by exome sequencing

Two groups of patients (n=58) were referred to exome sequencing. Group 1 were the 48 patients with inconclusive or negative genotypes tested by multiplex PCR (Figure 1). Group 2 consisted of 10 patients with either homozygous or compound heterozygous for NM_004004.6:c.109G>A in the *GJB2* gene. In order to exclude other potential molecular etiologies, these 10 patients were referred to exome sequencing owing to variable expressivity and incomplete penetration of NM_004004.6:c.109G>A in the *GJB2* gene.^20^

Of 48 patients from group 1, exome sequencing identified eleven diagnoses (23%, 11/48) and four probably diagnoses (8%, 4/48) (Table 3). No other causing variants related to hearing loss were identified from group 2. It was worth noting that the patient P27 was first identified as homozygous for NM_001038603.3(*MARVELD2*):c.1208_1211delGACA by exome sequencing. Considering a non-consanguineous family history, we performed qPCR and recovered a deletion covering exon 3 to exon 5 in the *MARVELD2* gene. Variant c.1208_1211delGACA locates in exon 4, where the other allele was deleted. Thus, we can conclude that c.1208_1211delGACA is strictly hemizygous in this case.

**Table 3.**
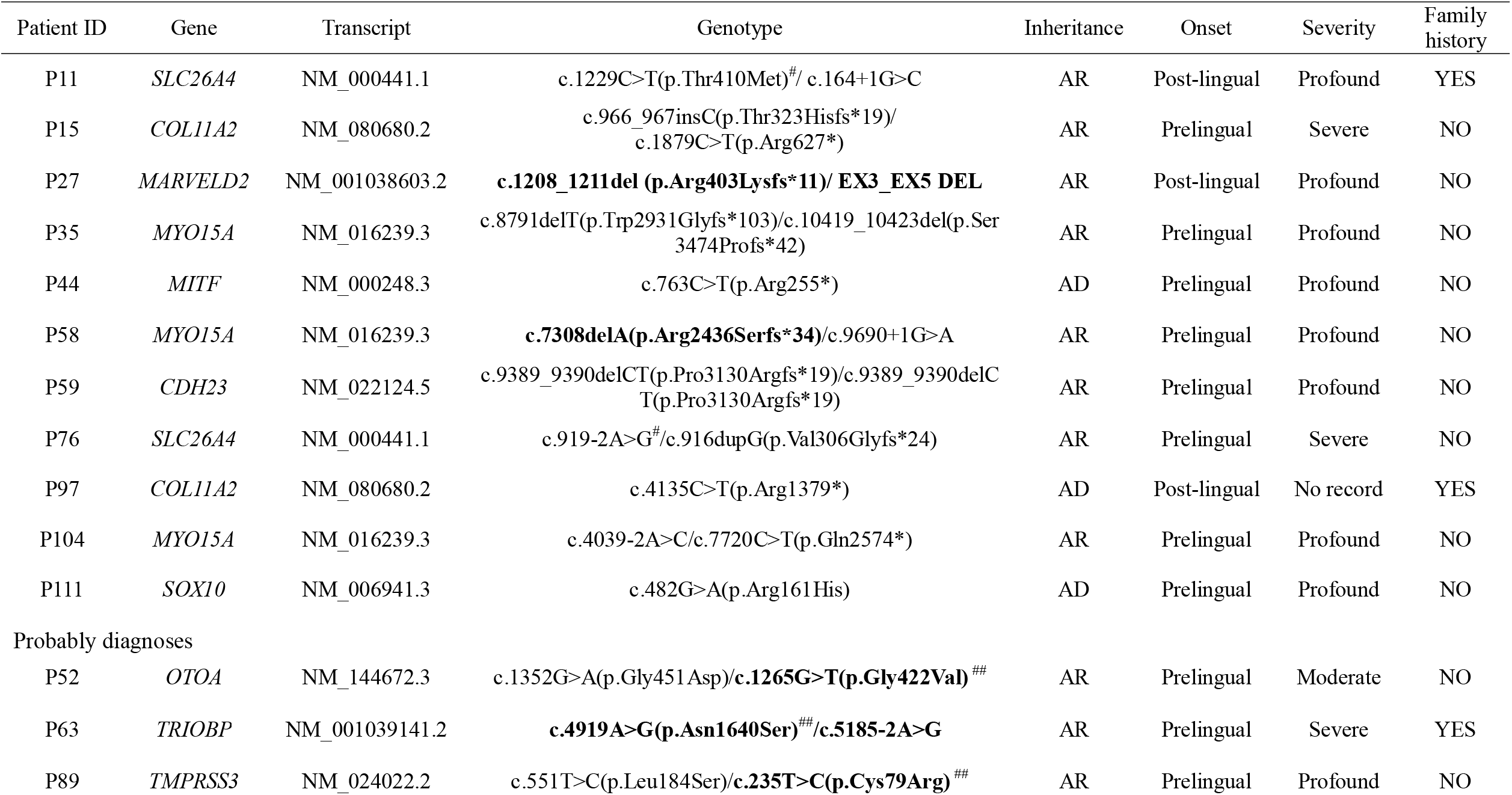
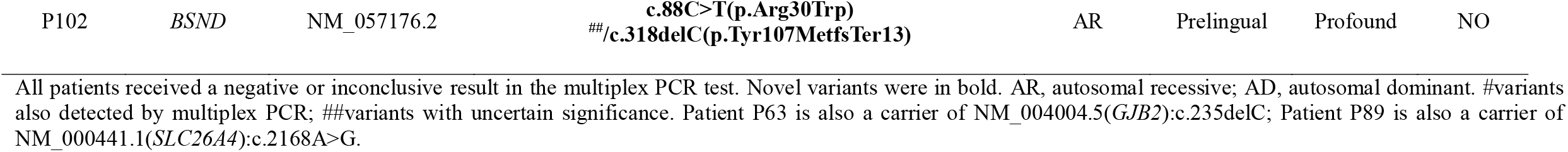
Diagnoses solely made by exome sequencing.

All patients received a negative or inconclusive result in the multiplex PCR test. Novel variants were in bold. AR, autosomal recessive; AD, autosomal dominant. #variants also detected by multiplex PCR; ##variants with uncertain significance. Patient P63 is also a carrier of NM_004004.5(*GJB2*):c.235delC; Patient P89 is also a carrier of NM_000441.1(*SLC26A4*):c.2168A>G.

To conclude, the 2-step approach identified the molecular etiology of 59/92 (64%) non-syndromic hearing loss index patients, including 44 (48%) diagnoses by multiplex PCR in the first step and 15 (16%) diagnoses/probable diagnoses by exome sequencing (Figure 2). Mutations in *GJB2* were most frequently detected (27/59), followed by mutations in *SLC26A4* (18/59), *MYO15A* (3/59), *COL11A2* (2/59), *MT-RNR1, MARVELD2, MITF, CDH23, OTOA, TRIOBP, TMPRSS3, SOX10* and *BSND* (1/59 each).

**Figure 2.**
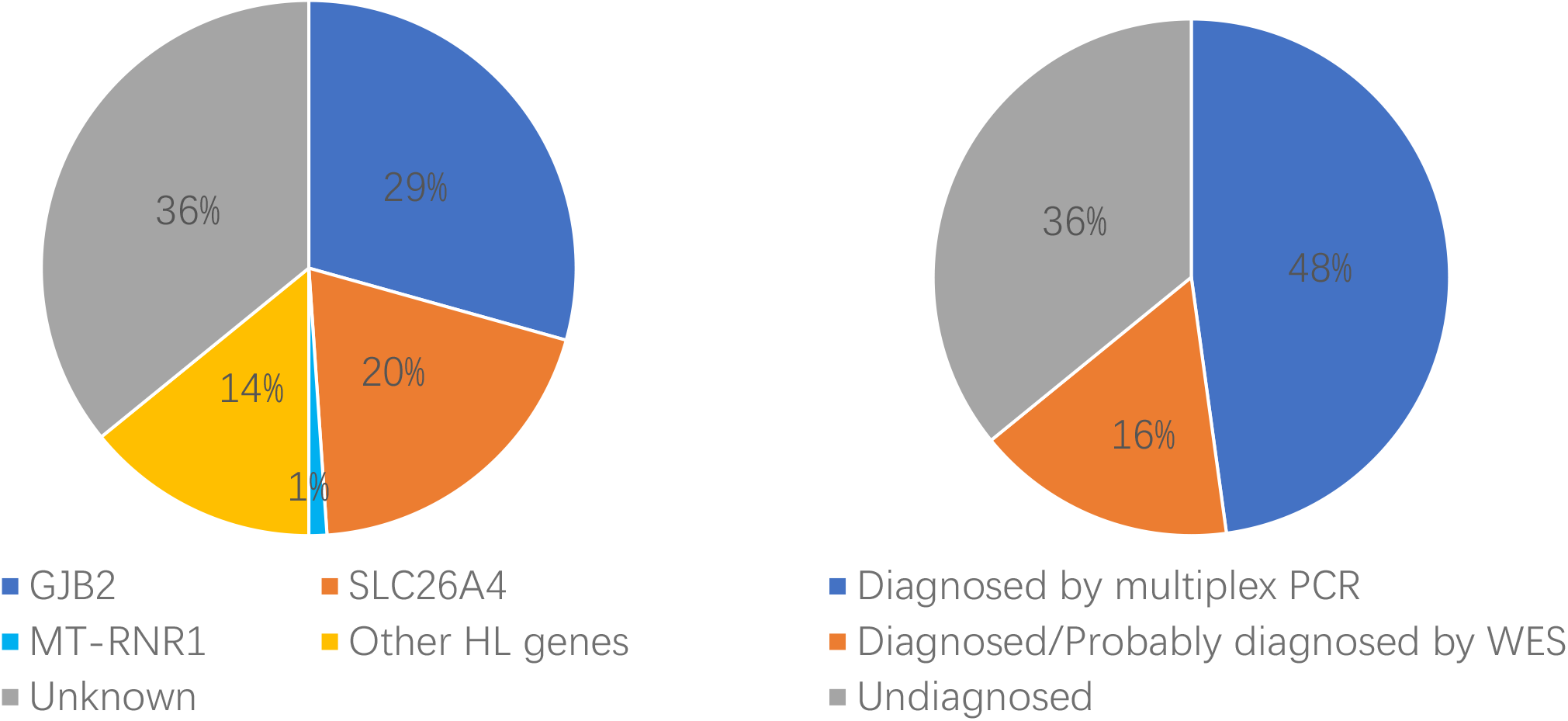
Genetic spectrum of enrolled non-syndromic hearing loss probands. Molecular diagnostic results were classified according to genes and detection methods.

Additionally, analysis of the exome sequencing data to discover secondary findings was done. The cohort displayed two pathogenic variants among the 59 secondary findings genes recommended by the ACMG^21^ (Supplementary table 1). *GLA*(NM_000169.3):c.1067G>A was identified in patient P21. It is related to Fabry disease, an X-linked inborn error of glycosphingolipid catabolism resulting from deficient or absent activity of the lysosomal enzyme alpha-galactosidase A.^22^ The second pathogenic variant was *RYR1*(NM_000540.3):c.6502G>A, may causing malignant hyperthermia. The rate of secondary findings in our cohort is 3.4% (2/58), comparable to the rate of previously published as 1.8% to 4.6%.^23-26^

## Discussion

This study applied a tiered, genetic testing approach to explore molecular diagnoses for a non-syndromic hearing loss cohort, achieving 64% (59/92) diagnostic yield. Although the diagnostic yield varies in different patient cohorts and depends on detection methods, our diagnostic yield is comparable to a multi-ethnic cohort tested using exome sequencing.^11^

This study addressed the etiology of 44 indexes in the first tier by using a commercial multiplex PCR kit, designing a rapid molecular diagnosis and saving the cost of exome sequencing. The multiplex PCR contains amplicons covering *GJB2, SLC26A4* and *MT-RNR1* genes. These three genes are known to have hotspot variants causing non-syndromic hearing loss in Asian populations.^27 28^ Compared to a single gene test of *GJB2*, which is frequently used as the first-tier test to exclude hotspot variants before exome sequencing,^7 11^ a multiplex PCR sequencing approach appears to be efficient and cost-effective as it is more flexible to detect hotspot variants across multiple deafness-related genes.

The allelic heterogenicity is common in hearing loss and associated with clinical phenotype heterogeneity, with both syndromic and nonsyndromic hearing loss being caused by mutations within the same gene.^29^ In this study, although we only enrolled patients with nonsyndromic hearing loss, deafness-related variants were identified in syndromic genes. P44 was heterozygous with a disease-causing nonsense variant in the *MITF* gene and P111 was heterozygous with a disease-causing missense variant in the *SOX10* gene. Both were associated with autosomal dominant inherited Waardenburg syndrome, which is called non-syndromic hearing loss mimics. The variability of phenotype challenges the clinical diagnosis and variant interpretation in genetic hearing loss.^18^

The molecular diagnosis of nonsyndromic hearing loss is challenged by the variable expressivity and high prevalence of NM_004004.6:c.109G>A in the *GJB2* gene.^20^ In our cohort, one patient with enlarged vestibular aqueduct was homozygous for both NM_000441.2:c.919-2A>G in the *SLC26A4* gene and NM_004004.6:c.109G>A in the *GJB2* gene in the first-tier. The genotype-phenotype consistency led us to determine that NM_000441.2:c.919-2A>G in the *SLC26A4* gene was the disease-causing variant. On the other hand, 10 patients solely diagnosed with NM_004004.6:c.109G>A in the *GJB2* gene in the first-tier, were referred to a further exome sequencing. No other potential molecular explanations were identified. These results indicate the importance of incorporating phenotype and genotype in practice.

Copy number variants are a common cause of nonsyndromic hearing loss.^30^ Exome sequencing data is feasible to analyze CNV, although it is characterized by low sensitivity and uncertain specificity.^31^ In this study, one patient in our cohort was diagnosed by a SNV compound with a CNV in the *MARVELD2*. The homozygous of c.1208_1211delGACA in the *MARVELD2* gene was initially thought as the causing etiology. The lack of consanguineous history led us to reanalyze the cover depth of exons in the *MARVELD2* gene, and identified an EX3_EX5 Del. It presented the importance to question a CNV deletion in a non-consanguineous family when a pathogenic variant was identified in a homozygous state.

Worthy of note, 17% (16/92) of patients passed the “newborn hearing screening” at birth but developed hearing loss at a later stage. Of these 16 patients, seven were molecularly diagnosed, spanning variants in the *GJB2* and *SLC26A4* genes (Supplementary Table S2). The results were consistent with the recent findings that newborns with positive genotypes could be missed by physiologic newborn hearing screens but identified by genetic screens, highlighting the necessity of concurrent hearing and genetic screening in newborns.^28 32^

In conclusion, this work demonstrates the benefits of a stepwise approach to diagnose non-syndromic hearing loss patients. Instead of using exome sequencing in the first beginning, multiplex PCR targeted hotspot variants across multiple genes can provide molecular etiology for 48% of patients in a promptly and effective manner. A further cost-effectiveness analysis is assured.

## Data Availability

All data relevant to the study are included in the article or uploaded as supplementary information.

## Acknowledgements

The authors thank the patients for their participation.

## Contributors

ZP conceived and designed the study. LC, JW and JX contributed to patient recruitment and phenotypic information collection. JP performed bioinformatics analysis. HL, XX, NL, and CC performed the validation experiments. XX, NL, CC, LC and NS performed data analysis and interpretation. JW, JX and LC drafted the manuscript. All authors read and approved the final manuscript.

## Funding

This work was supported by the Special Foundation for High-level Talents of Guangdong (grant 2016TX03R171) to Dr Peng. The funding sources had no role in the design and conduct of the study; collection, management, analysis, and interpretation of the data; preparation, review, or approval of the manuscript; and decision to submit the manuscript for publication.

## Competing interests

JX, LC, HL, JP, NS, and ZP were employed at BGI Genomics at the time of submission. No other conflicts relevant to this study should be reported.

## Patient consent for publication

Not required.

## Provenance and peer review

Not commissioned; externally peer reviewed.

## Notes

### Author Declarations

Institutional Review Board of BGI

## Reference

1. Morton CC, Nance WE. Newborn hearing screening--a silent revolution. N Engl J Med 2006;354(20):2151–64.

2. Morton CCN, Walter E. Newborn hearing screening--a silent revolution. The New England Journal of Medicine 2006;354(20):2151–64.

3. Vona B, Nanda I, Hofrichter MA, Shehata-Dieler W, Haaf T. Non-syndromic hearing loss gene identification: A brief history and glimpse into the future. Mol Cell Probes 2015;29(5):260–70.

4. Van Camp G, Smith R. Hereditary Hearing Loss Homepage. https://hereditaryhearingloss.org. Accessed: August 2019.

5. Li M, Mei L, He C, Chen H, Cai X, Liu Y, Tian R, Tian Q, Song J, Jiang L, Liu C, Wu H, Li T, Liu J, Li X, Yi Y, Yan D, Blanton SH, Hu Z, Liu X, Li J, Ling J, Feng Y. Extrusion pump ABCC1 was first linked with nonsyndromic hearing loss in humans by stepwise genetic analysis. Genet Med 2019.

6. Korver AMH, Smith RJH, Van Camp G, Schleiss MR, Bitner-Glindzicz MAK, Lustig LR, Usami S-i, Boudewyns AN. Congenital hearing loss. Nature Reviews Disease Primers 2017;3(1).

7. Guan Q, Balciuniene J, Cao K, Fan Z, Biswas S, Wilkens A, Gallo DJ, Bedoukian E, Tarpinian J, Jayaraman P, Sarmady M, Dulik M, Santani A, Spinner N, Abou Tayoun AN, Krantz ID, Conlin LK, Luo M. AUDIOME: a tiered exome sequencing-based comprehensive gene panel for the diagnosis of heterogeneous nonsyndromic sensorineural hearing loss. Genet Med 2018;20(12):1600–08.

8. Almontashiri NAM, Alswaid A, Oza A, Al-Mazrou KA, Elrehim O, Tayoun AA, Rehm HL, Amr SS. Recurrent variants in OTOF are significant contributors to prelingual nonsydromic hearing loss in Saudi patients. Genet Med 2018;20(5):536–44.

9. Sloan-Heggen CM, Bierer AO, Shearer AE, Kolbe DL, Nishimura CJ, Frees KL, Ephraim SS, Shibata SB, Booth KT, Campbell CA, Ranum PT, Weaver AE, Black-Ziegelbein EA, Wang D, Azaiez H, Smith RJH. Comprehensive genetic testing in the clinical evaluation of 1119 patients with hearing loss. Hum Genet 2016;135(4):441–50.

10. Norris VW, Arnos KS, Hanks WD, Xia X, Nance WE, Pandya A. Does universal newborn hearing screening identify all children with GJB2 (Connexin 26) deafness? Penetrance of GJB2 deafness. Ear Hear 2006;27(6):732–41.

11. Bademci G, Foster J, 2nd, Mahdieh N, Bonyadi M, Duman D, Cengiz FB, Menendez I, Diaz-Horta O, Shirkavand A, Zeinali S, Subasioglu A, Tokgoz-Yilmaz S, Huesca-Hernandez F, de la Luz Arenas-Sordo M, Dominguez-Aburto J, Hernandez-Zamora E, Montenegro P, Paredes R, Moreta G, Vinueza R, Villegas F, Mendoza-Benitez S, Guo S, Bozan N, Tos T, Incesulu A, Sennaroglu G, Blanton SH, Ozturkmen-Akay H, Yildirim-Baylan M, Tekin M. Comprehensive analysis via exome sequencing uncovers genetic etiology in autosomal recessive nonsyndromic deafness in a large multiethnic cohort. Genet Med 2016;18(4):364–71.

12. Zazo Seco C, Wesdorp M, Feenstra I, Pfundt R, Hehir-Kwa JY, Lelieveld SH, Castelein S, Gilissen C, de Wijs IJ, Admiraal RJ, Pennings RJ, Kunst HP, van de Kamp JM, Tamminga S, Houweling AC, Plomp AS, Maas SM, de Koning Gans PA, Kant SG, de Geus CM, Frints SG, Vanhoutte EK, van Dooren MF, van den Boogaard MH, Scheffer H, Nelen M, Kremer H, Hoefsloot L, Schraders M, Yntema HG. The diagnostic yield of whole-exome sequencing targeting a gene panel for hearing impairment in The Netherlands. Eur J Hum Genet 2017;25(3):308–14.

13. Baux D, Vache C, Blanchet C, Willems M, Baudoin C, Moclyn M, Faugere V, Touraine R, Isidor B, Dupin-Deguine D, Nizon M, Vincent M, Mercier S, Calais C, Garcia-Garcia G, Azher Z, Lambert L, Perdomo-Trujillo Y, Giuliano F, Claustres M, Koenig M, Mondain M, Roux AF. Combined genetic approaches yield a 48% diagnostic rate in a large cohort of French hearing-impaired patients. Sci Rep 2017;7(1):16783.

14. Budde BS, Aly MA, Mohamed MR, Bress A, Altmuller J, Motameny S, Kawalia A, Thiele H, Konrad K, Becker C, Toliat MR, Nurnberg G, Sayed EAF, Mohamed ES, Pfister M, Nurnberg P. Comprehensive molecular analysis of 61 Egyptian families with hereditary nonsyndromic hearing loss. Clin Genet 2020.

15. Huang J, Liang X, Xuan Y, Geng C, Li Y, Lu H, Qu S, Mei X, Chen H, Yu T, Sun N, Rao J, Wang J, Zhang W, Chen Y, Liao S, Jiang H, Liu X, Yang Z, Mu F, Gao S. A reference human genome dataset of the BGISEQ-500 sequencer. Gigascience 2017;6(5):1–9.

16. DePristo MA, Banks E, Poplin R, Garimella KV, Maguire JR, Hartl C, Philippakis AA, del Angel G, Rivas MA, Hanna M, McKenna A, Fennell TJ, Kernytsky AM, Sivachenko AY, Cibulskis K, Gabriel SB, Altshuler D, Daly MJ. A framework for variation discovery and genotyping using next-generation DNA sequencing data. Nat Genet 2011;43(5):491–8.

17. DiStefano MT, Hemphill SE, Oza AM, Siegert RK, Grant AR, Hughes MY, Cushman BJ, Azaiez H, Booth KT, Chapin A, Duzkale H, Matsunaga T, Shen J, Zhang W, Kenna M, Schimmenti LA, Tekin M, Rehm HL, Tayoun ANA, Amr SS. ClinGen expert clinical validity curation of 164 hearing loss gene–disease pairs. Genetics in Medicine 2019.

18. Oza AM, DiStefano MT, Hemphill SE, Cushman BJ, Grant AR, Siegert RK, Shen J, Chapin A, Boczek NJ, Schimmenti LA, Murry JB, Hasadsri L, Nara K, Kenna M, Booth KT, Azaiez H, Griffith A, Avraham KB, Kremer H, Rehm HL, Amr SS, Abou Tayoun AN, ClinGen Hearing Loss Clinical Domain Working G. Expert specification of the ACMG/AMP variant interpretation guidelines for genetic hearing loss. Hum Mutat 2018;39(11):1593–613.

19. Rosenbloom KR, Armstrong J, Barber GP, Casper J, Clawson H, Diekhans M, Dreszer TR, Fujita PA, Guruvadoo L, Haeussler M, Harte RA, Heitner S, Hickey G, Hinrichs AS, Hubley R, Karolchik D, Learned K, Lee BT, Li CH, Miga KH, Nguyen N, Paten B, Raney BJ, Smit AF, Speir ML, Zweig AS, Haussler D, Kuhn RM, Kent WJ. The UCSC Genome Browser database: 2015 update. Nucleic Acids Res 2015;43(Database issue):D670–81.

20. Shen J, Oza AM, Del Castillo I, Duzkale H, Matsunaga T, Pandya A, Kang HP, Mar-Heyming R, Guha S, Moyer K, Lo C, Kenna M, Alexander JJ, Zhang Y, Hirsch Y, Luo M, Cao Y, Wai Choy K, Cheng YF, Avraham KB, Hu X, Garrido G, Moreno-Pelayo MA, Greinwald J, Zhang K, Zeng Y, Brownstein Z, Basel-Salmon L, Davidov B, Frydman M, Weiden T, Nagan N, Willis A, Hemphill SE, Grant AR, Siegert RK, DiStefano MT, Amr SS, Rehm HL, Abou Tayoun AN, ClinGen Hearing Loss Working G. Consensus interpretation of the p.Met34Thr and p.Val37Ile variants in GJB2 by the ClinGen Hearing Loss Expert Panel. Genet Med 2019;21(11):2442–52.

21. Green RC, Berg JS, Grody WW, Kalia SS, Korf BR, Martin CL, McGuire AL, Nussbaum RL, O’Daniel JM, Ormond KE, Rehm HL, Watson MS, Williams MS, Biesecker LG, American College of Medical G, Genomics. ACMG recommendations for reporting of incidental findings in clinical exome and genome sequencing. Genet Med 2013;15(7):565–74.

22. Schiffmann R. Fabry disease. Neurocutaneous Syndromes2015:231–48.

23. Yang Y, Muzny DM, Xia F, Niu Z, Person R, Ding Y, Ward P, Braxton A, Wang M, Buhay C, Veeraraghavan N, Hawes A, Chiang T, Leduc M, Beuten J, Zhang J, He W, Scull J, Willis A, Landsverk M, Craigen WJ, Bekheirnia MR, Stray-Pedersen A, Liu P, Wen S, Alcaraz W, Cui H, Walkiewicz M, Reid J, Bainbridge M, Patel A, Boerwinkle E, Beaudet AL, Lupski JR, Plon SE, Gibbs RA, Eng CM. Molecular findings among patients referred for clinical whole-exome sequencing. JAMA 2014;312(18):1870–9.

24. Schwartz MLB, McCormick CZ, Lazzeri AL, Lindbuchler DM, Hallquist MLG, Manickam K, Buchanan AH, Rahm AK, Giovanni MA, Frisbie L, Flansburg CN, Davis FD, Sturm AC, Nicastro C, Lebo MS, Mason-Suares H, Mahanta LM, Carey DJ, Williams JL, Williams MS, Ledbetter DH, Faucett WA, Murray MF. A Model for Genome-First Care: Returning Secondary Genomic Findings to Participants and Their Healthcare Providers in a Large Research Cohort. Am J Hum Genet 2018;103(3):328–37.

25. Dewey FE, Murray MF, Overton JD, Habegger L, Leader JB, Fetterolf SN, O’Dushlaine C, Van Hout CV, Staples J, Gonzaga-Jauregui C, Metpally R, Pendergrass SA, Giovanni MA, Kirchner HL, Balasubramanian S, Abul-Husn NS, Hartzel DN, Lavage DR, Kost KA, Packer JS, Lopez AE, Penn J, Mukherjee S, Gosalia N, Kanagaraj M, Li AH, Mitnaul LJ, Adams LJ, Person TN, Praveen K, Marcketta A, Lebo MS, Austin-Tse CA, Mason-Suares HM, Bruse S, Mellis S, Phillips R, Stahl N, Murphy A, Economides A, Skelding KA, Still CD, Elmore JR, Borecki IB, Yancopoulos GD, Davis FD, Faucett WA, Gottesman O, Ritchie MD, Shuldiner AR, Reid JG, Ledbetter DH, Baras A, Carey DJ. Distribution and clinical impact of functional variants in 50,726 whole-exome sequences from the DiscovEHR study. Science 2016;354(6319).

26. Haer-Wigman L, van der Schoot V, Feenstra I, Vulto-van Silfhout AT, Gilissen C, Brunner HG, Vissers L, Yntema HG. 1 in 38 individuals at risk of a dominant medically actionable disease. Eur J Hum Genet 2019;27(2):325–30.

27. Tsukada K, Nishio SY, Hattori M, Usami S. Ethnic-specific spectrum of GJB2 and SLC26A4 mutations: their origin and a literature review. Ann Otol Rhinol Laryngol 2015;124 Suppl 1:61S-76S.

28. Wang Q, Xiang J, Sun J, Yang Y, Guan J, Wang D, Song C, Guo L, Wang H, Chen Y, Leng J, Wang X, Zhang J, Han B, Zou J, Yan C, Zhao L, Luo H, Han Y, Yuan W, Zhang H, Wang W, Wang J, Yang H, Xu X, Yin Y, Morton CC, Zhao L, Zhu S, Shen J, Peng Z. Nationwide population genetic screening improves outcomes of newborn screening for hearing loss in China. Genet Med 2019;21(10):2231–38.

29. Keats BJ, Berlin CI. Genomics and hearing impairment. Genome Res 1999;9(1):7–16.

30. Shearer AE, Kolbe DL, Azaiez H, Sloan CM, Frees KL, Weaver AE, Clark ET, Nishimura CJ, Black-Ziegelbein EA, Smith RJ. Copy number variants are a common cause of non-syndromic hearing loss. Genome Med 2014;6(5):37.

31. Yao R, Zhang C, Yu T, Li N, Hu X, Wang X, Wang J, Shen Y. Evaluation of three read-depth based CNV detection tools using whole-exome sequencing data. Molecular Cytogenetics 2017;10(1).

32. Guo L, Xiang J, Sun L, Yan X, Yang J, Wu H, Guo K, Peng J, Xie X, Yin Y, Wang J, Yang H, Shen J, Zhao L, Peng Z. Concurrent hearing and genetic screening in a general newborn population. Hum Genet 2020;139(4):521–30.

